# A Multi-Centre, Tolerability Study of a Cannabidiol-enriched Cannabis Herbal Extract for Chronic Headaches in Adolescents: the CAN-CHA Protocol

**DOI:** 10.1101/2023.08.04.23293647

**Authors:** Manik Chhabra, Evan C. Lewis, Robert Balshaw, Breanne Stewart, Zina Zaslawski, Trinity Lowthian, Zahra Alidina, Melila Chesick-Gordis, Wenli Xie, Britt I. Drögemöller, Galen E.B. Wright, Kathryn A Birnie, Katelynn E Boerner, Vivian W. L. Tsang, Samantha Lee Irwin, Daniela Pohl, Alexander G Weil, Erick Sell, Erika Penz, Amy Robson- MacKay, Sophia Mbabaali, Stephanie Blackman, Shanlea Gordon, Jane Alcorn, Richard J. Huntsman, Tim F Oberlander, G Allen Finley, Lauren E Kelly

## Abstract

**Introduction:** Cannabis products have been used in the management of headaches in adults and may play a role in pediatric chronic pain. Canadian pediatricians report increasing use of cannabis for the management of chronic headaches, despite no well-controlled studies to inform its dosing, safety, and effectiveness. The aim of our clinical trial is to determine the dosing and safety of a Cannabidiol (CBD)-enriched Cannabis Herbal Extract (CHE) for the treatment of chronic headaches in adolescents.

**Methods and analysis:** Youth, parents and an expert steering committee co-designed this tolerability study. Twenty adolescents (aged 14 to 17 years), with a chronic migraine diagnosis for more than 6 months that has not responded to other therapies, will be enrolled into an open label, dose escalation study across three Canadian sites. Study participants will receive escalating doses of a CBD-enriched CHE (MPL-001 with a THC:CBD of 1:25), starting at 0.2-0.4 mg/kg of CBD per day escalating monthly up to 0.8-1.0 mg/kg of CBD per day. The primary objective of this study is to determine the safety and tolerability of CBD-enriched CHE in adolescents with chronic migraine. Secondary objectives of this study will inform the development of subsequent randomized controlled trials and include investigating the relationship between the dose escalation and change in the frequency of headache, impact and intensity of pain, changes in sleep, mood, function, and quality of life. Exploratory outcomes include investigating steady-state trough plasma levels of bioactive cannabinoids and investigating how pharmacogenetic profiles affect cannabinoid metabolism among adolescents receiving CBD-enriched CHE.

**Discussion:** This protocol was co-designed with youth and describes a tolerability clinical trial of CBD-enriched CHE in adolescents with chronic headaches that have not responded to conventional therapies. This study is the first clinical trial on cannabis products in adolescents with chronic headaches and will inform the development of future comparative effectiveness clinical trials.

**Trial registration:** CAN-CHA trial is registered with ClinicalTrials.gov with a number of register NCT05337033.

## Introduction

Globally, chronic headaches are one of the major causes of disability among adolescents. The World Health Organization classifies it under the top ten disabling health conditions,[1, 2] with a prevalence of 7.8% in adolescents 14 years of age or older [3–5]. In the US alone, the total annual cost incurred by pediatric headache is estimated around $1.1 billion [6]. Adolescents with chronic headaches often experience reduced quality of life, sleep disruption, anxiety, fatigue, limb pain, dizziness, overuse of medications and academic challenges [7]. Despite advancements in the therapeutic management of chronic pain, treatment of chronic headache disorders in adolescents remains challenging. Non-Steroidal Anti-Inflammatory Drugs (NSAIDs), triptans, gabapentin, neuromodulation devices and ergotamine are used for the acute treatment of migraine [8, 9]. Preventive migraine therapies are limited in availability, efficacy, and authorization for use in the adolescent population. Topiramate is the only FDA approved preventive treatment for migraine in adolescents [1, 10, 11]. In other areas of medicine, cannabinoids have shown therapeutic potential in adolescents where conventional medications fail, including treatment resistant epilepsy, and chemotherapy-induced nausea and vomiting [12]. In adults, use of cannabis products is increasing for the treatment of headaches and migraines [13]. An observational study reported that 36% of adult cannabis users are using cannabis to relieve symptoms related to migraine and/or headaches. Further, this study reported that the use of cannabis products in adults led to an average reduction of 3.6 points on a 10-point intensity of headache scale [14, 15]. Despite promising observational data in adults, a paucity of literature exists that demonstrates the tolerability of cannabis for the treatment of chronic headaches in adolescents. Cannabidiol (CBD) and Tetrahydrocannabinol (THC) are the principal active cannabinoids which have a number of potential therapeutic applications [16]. CBD, which is not associated with the same intoxicating effects of THC, acts as a negative allosteric modulator of CB1 receptors in the endocannabinoid system [17]. CBD potentiates anandamide-mediated intrinsic neurotransmission [18], and has antioxidant and anti-inflammatory activity [19].

Canadian studies have demonstrated the safety and tolerability of CBD-enriched CHE in children with refractory epilepsy [20, 21]. Huntsman et al reported on the preliminary results of the CARE-E trial where a 1:20 THC:CBD CHE oil was found to be well tolerated in children, and THC plasma concentrations were below levels associated with intoxication despite CBD doses of up to 12 mg/kg per day [21].

In Canada, recreational markets have increased cannabis accessibility and there is increasing interest in managing chronic headaches off-label with cannabinoids, self-guided in the absence of evidence [22]. The paucity of clinical trial data in children is due to many historical challenges with studying cannabis products and cannabinoids, resulting from legal difficulties in obtaining products, variable product quality control and the stigma associated with illegal drug use, and particularly with children. The current reality warrants the need to conduct robust interventional studies establishing the tolerability, safety, and efficacy of cannabis in adolescents with chronic headaches. Here we describe a protocol for CAN-CHA (<u>CANnabis for Chronic Headaches in Adolescents)</u> trial, an open-label dose escalation study to establish the tolerability of a CBD-enriched Cannabis Herbal Extract (CHE) in adolescents with chronic headaches. CAN-CHA is funded by the SickKids Foundation, sponsored by the University of Manitoba and has received approvals from Health Canada and research ethics board. CAN-CHA is registered with ClinicalTrials.gov (NCT05337033).

## Methods and analysis

### Primary Objective

To determine the safety and tolerability of escalating doses of a CBD-enriched CHE in adolescents with chronic headaches.

### Secondary Objectives

1. To investigate the relationship between the dose-escalation with headache-free days.
2. To monitor the effect of CBD-enriched CHE oil on the intensity of pain related to chronic headaches.
3. To evaluate the effect of CBD-enriched CHE oil on sleep, mood, and function in adolescents with chronic headaches.
4. To explore the impact of chronic headaches on quality of life.

### Exploratory Objectives

1. To investigate the relationship between the dose-escalation and steady-state trough levels of bioactive cannabinoids/endocannabinoids.
2. To study pharmacogenetic variations among adolescents receiving CBD-enriched CHE oil.

### Study Population

We will recruit 20 adolescents across three study sites: Halifax, Toronto, and Vancouver. To be eligible to participate in this study, an individual must meet all of the following criteria:

1. Adolescents between 14-17 years of age at the time of screening
2. Diagnosed with Chronic Migraine according to ICHD-3: headache (migraine-like or tension-type like) occurring on 15 or more days per month for more than 3 months, which on at least 8 days per month have features of migraine headache [23].
3. Failed at least two preventive treatment options on the grounds of tolerability and/or efficacy, including <u>but not limited</u> to antidepressants (tricyclic antidepressant or selective norepinephrine reuptake inhibitor), magnesium, gabapentin, topiramate, beta-blockers, memantine, and/or non-pharmacological therapies including nutraceuticals and botox.
4. Females who have reached menarche must have a negative serum pregnancy test during screening
5. Must be willing to engage with psychology and physiotherapy throughout the trial as appropriate.

Adolescents meeting any of the following criteria will be <u>excluded</u> from the study:

1. As per the investigator judgment, the participant is not an ideal candidate due to a personal issue or medical condition that is likely to impede in the successful completion of the study
2. Participants with a history of post-concussion headache or new daily persistent headache
3. Participants with a diagnosis of medication overuse headache
4. Participants with cardiac, renal, or hepatic disease (assessed by the site investigator)
5. Participants with complex regional pain syndrome-II
6. Participants with abnormal ECG findings at baseline (as determined by the investigator)
7. Participants who are on the following medications: opioids, antipsychotics, antimanic, barbiturates, benzodiazepines, muscle relaxants, sedatives, or tramadol
8. Participants with developmental delay or impairments including autism, cerebral palsy, or intellectual disability
9. Participants with a personal or family history of schizophrenia or psychotic disorders
10. Participants who are/plan to become pregnant within the study period or within three months of interventional product discontinuation
11. Participants who cannot commit to using contraception, refraining from recreational cannabis use and driving throughout the study period

## Withdrawal criteria

Study participants may withdraw from the study at any point. If a participant’s headaches worsen or they suffer from intolerable treatment related adverse effects, they will be withdrawn from the study. Participants who become pregnant during the study period, do not attend follow-up visits, or do not comply with the prescribed interventional drug regimen will be withdrawn from the study. Participants will be given the option to only withdraw from the study intervention, these participants will continue to be followed up for safety assessments till the end of study. All participants withdrawn from the study will be included in the final report at the end of the study for transparency.

## Intervention

The investigational product for our study is a CBD-enriched CHE, MPL-001, purchased from MediPharm Labs. MPL-001 is a CBD-enriched CHE where each ml of oil contains 2 mg of THC and 50 mg of CBD dissolved in coconut/palm-based medium chain triglycerides (MCT) carrier oil. The MPL-001 (CBD:THC 25:1) oil used in this study contains lemon-peppermint flavoring agents. Manufacturing of MPL-001 occurs following Good Manufacturing Practices includes the following steps: harvesting of plant, followed by weighting, and drying. Further, dried plants are subjected to bucking, ethanolic extraction, filtration of cannabinoid ethanolic solution, and evaporation of ethanol, this leaves over acidic cannabis resin. Subsequently, cannabis resin is decarboxylated (activated) and mixed with oil. The oil is made available in a glass bottle sealed with child lock caps, stored and labeled at the study central pharmacy according in accordance with the *Cannabis Act, 2018* and the Health Canada Division 5 *Food and Drugs Act* [24]. All the study participants will receive an escalating dose starting at 0.2-0.4 mg/kg of CBD per day with dose increases (0.2 mg/kg/day increments) happening monthly to a maximum of 0.8-1.0 mg/kg of CBD per day. This dosing strategy reflects the current clinical practice of study investigators and well below the maximum dose of CBD-enriched CHE of 10-12 mg/kg/day previously well tolerated in children with epilepsy [21]. Participants will be provided with dosing calendars (Fig 2) and be instructed to take their daily dose bid with 25% of the dose in the morning and 75% of the daily dose in the late afternoon to mimic the diurnal variation [25] in the endocannabinoids and prevents adolescents from having to take cannabis during school hours.

## Study design

CAN-CHA is a multicenter, open-label dose-escalation study to determine the tolerability of MPL-001 in adolescents with treatment refractory chronic migraine. A traditional 3+3 dose finding design was not practical as all participants would not receive the lowest cannabinoid dose and the recommended “start low and go slow” approach to titration. The trial will be conducted in three Canadian centers. CAN-CHA will consist of three different phases: baseline (1 month) without intervention, treatment (4 months of escalating doses) and weaning (1 month). All participants and their caregivers will be invited to complete a pre– and post-study survey about their experiences in the trial to inform future research.

## Patient and public involvement

CAN-CHA trial was designed in collaboration with youth from the KidsCAN Young Persons’ Research Advisory Group (YRPAG) and from the Solutions for Kids in Pain (SKIP) network. The Canadian Collaborative for Childhood Cannabinoid Therapeutics (C4T) Parent Advisory Committee provided insight on the outcome measurement tools and the consent form. Three youth advisors (TL, ZA, MC-G) with chronic migraines have been involved throughout the study design process and will continue to advise on recruitment strategies, designing materials, reporting and dissemination.

## Baseline Phase

Eligible adolescents will be asked by their healthcare providers if they are interested in learning more about this research study. A study team member, who is not involved in the patients’ care, will present the study, review the consent documents and answer any questions from the patients and their families. Adolescents who meet the inclusion criteria, assent, and whose caregiver’s consent to participate will then begin a baseline period. Baseline will consist of one month period without intervention to estimate headache frequency and severity, as well as mood, sleep, and pain before administering the intervention to the participants. Participants will be asked to maintain a daily electronic headache diary that will include reporting on the severity of headaches, associated pain, sleep, absences from work/school and adverse events. Participants will complete age-validated scales on sleep related impairment, anxiety, depression, positive mood, pain interference, family impact and goal attainment scaling [26–31] as described in the Fig 1. A blood sample will be drawn from the study participants to measure liver transaminases (ALT/AST) and creatinine, measure endocannabinoids and detect pregnancies, while saliva will be sampled for extraction of genomic DNA to allow for genotyping of pharmacogenetic variants. ECGs (within 3 months of screening or done at the time of screening) will be recorded at baseline to ensure there are no cardiac electrical activity concerns.

**Fig 1.**
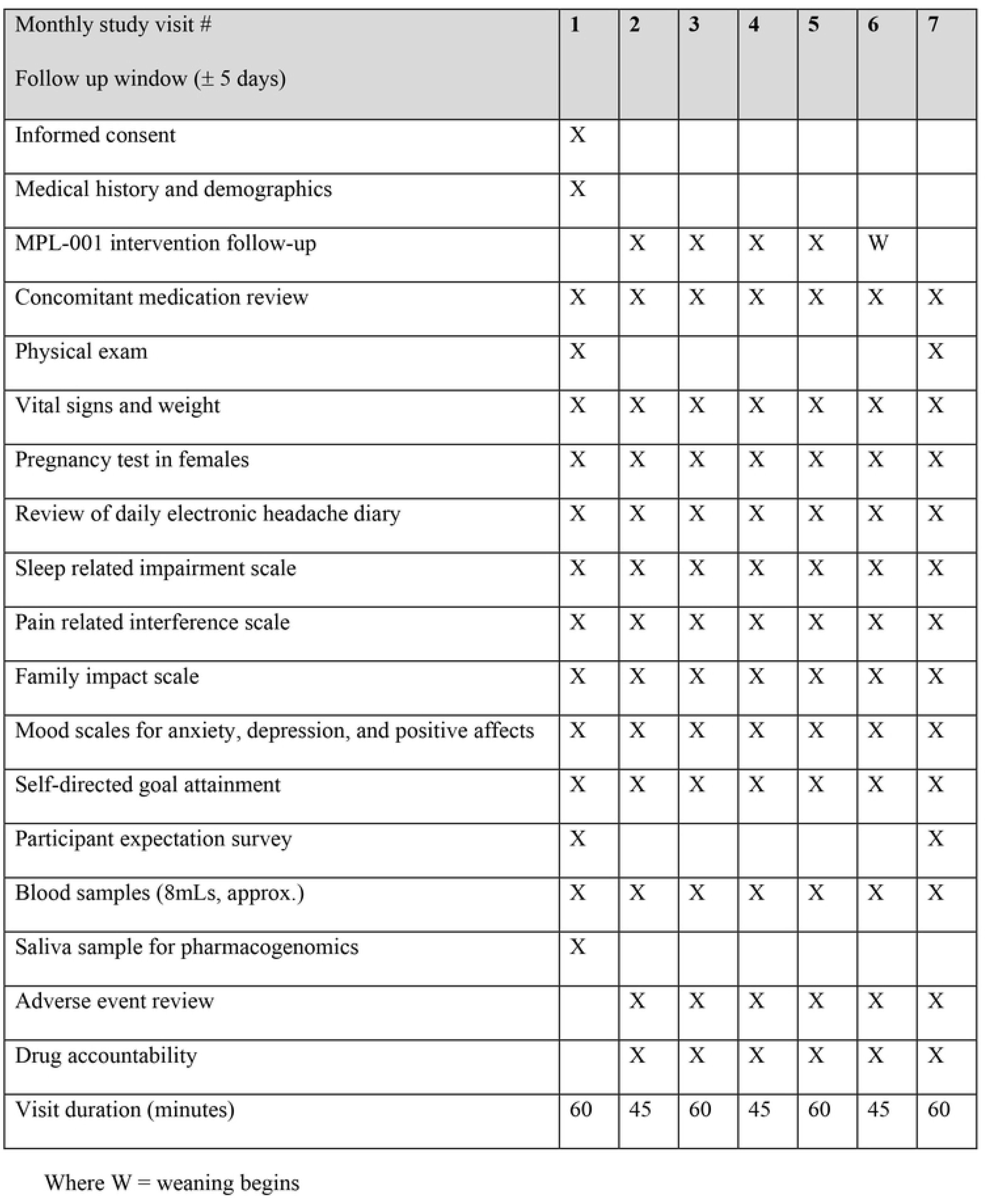
CAN-CHA schedule of events as per SPIRIT guidelines

**Fig 2.**
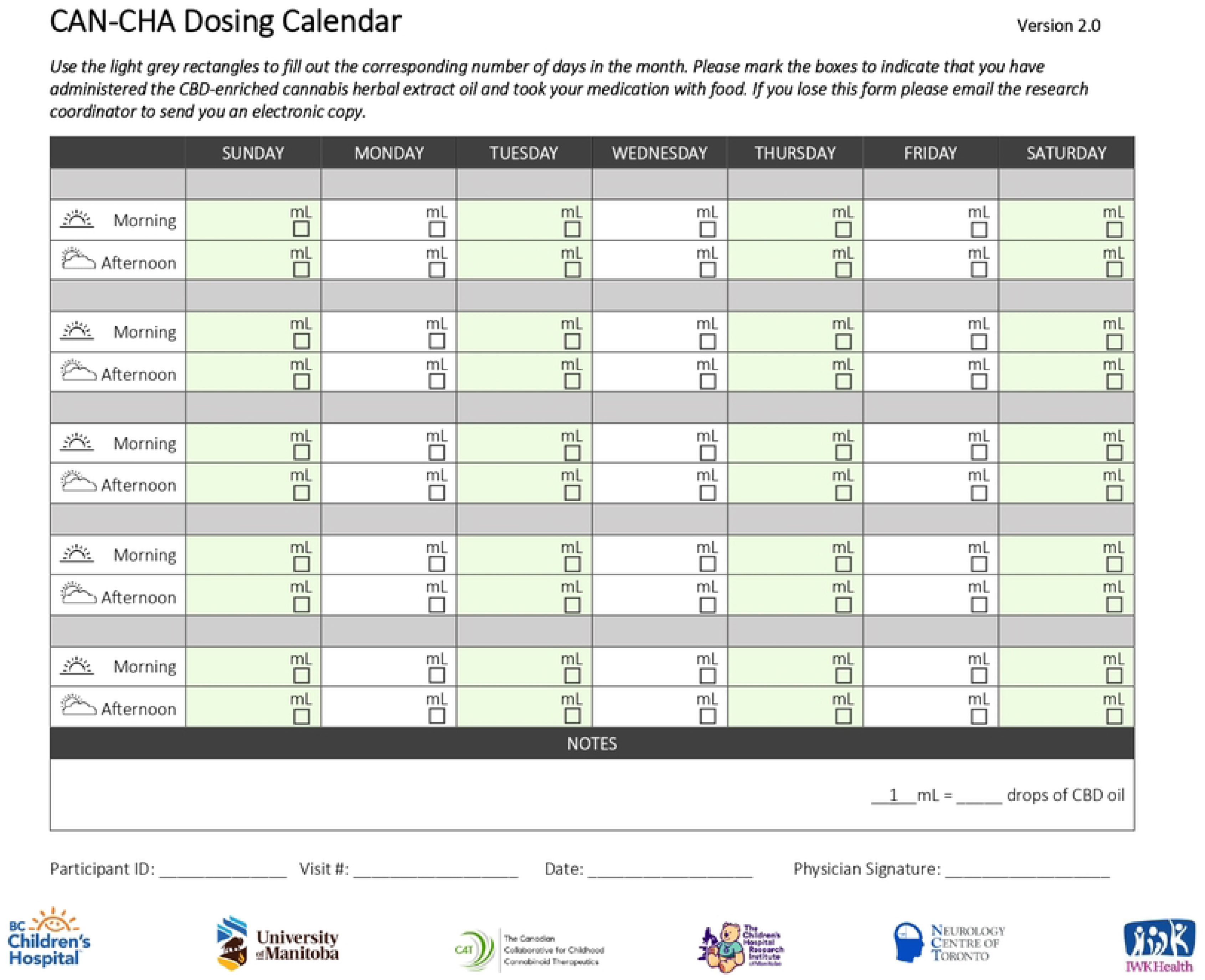
CAN-CHA dosing calendar.

## Treatment Phase

Following the one-month baseline period, study participants will receive CBD-enriched CHE oil, MPL-001with a dosing calendar and administration pamphlet shipped directly to their homes from the trial coordinating centre. A handout on cannabis oil administration co-created with the youth advisors and will be provided to all study participants and their families that will also have a video component. There will also be a demonstration by the research coordinator at the first study visit using olive oil in a product bottle. The study participants will be instructed to administer the investigational product at a starting dose of 0.2-0.4 mg/kg/day divided into two doses (BID, 25% in the morning and 75% in the evening after school) each day for one month.

Dose-escalation schedule during the treatment phase is described in Table 1. Participants will continue to complete a daily electronic headache diary to monitor symptoms and adverse effects. During all follow up visits, participants and their caregivers will be asked to complete validated outcome measurement tools alongside the PedsQL ™ Family Impact Module assessment, and Self-directed goal attainment. Blood samples will be collected prior to starting the next dose level, at study visits and will be used to evaluate changes and variability in cannabinoid pharmacokinetics with escalating doses, confirm pregnancies, and monitor liver enzymes and creatinine over time.

**Table 1.** Dosing schedule for CAN-CHA evaluating a CBD-enriched CHE from visit 2 to visit 6.

| Visit | CBD total daily dose | Frequency | Duration |
| --- | --- | --- | --- |
| Visit 2 | 0.2-0.4 mg/kg/day | BID | 1 month |
| Visit 3 | 0.4-0.6 mg/kg/day | BID | 1 month |
| Visit 4 | 0.6-0.8 mg/kg/day | BID | 1 month |
| Visit 5 | 0.8-1 mg/kg/day | BID | 1 month |

**Weaning Phase:** After the baseline phase (one month, no treatment) and the treatment phase (four months, escalating doses), participants will start the weaning schedule. Weaning includes incrementally reducing the dose (by 0.2 mg/kg CBD every week) leading to complete discontinuation of the study product. The complete weaning schedule is detailed in Table 2. The intervention will be discontinued by visit 7. If the parents, adolescents, and healthcare providers feel that there was improvement while on the intervention, participants will discuss the continued authorization of medical cannabis with the enrolling physician, caregivers and their healthcare team.

**Table 2.** The weaning schedule which begins at visit 6 and ends at visit 7.

| Visit 6 | CBD total daily dose | Frequency | Duration |
| --- | --- | --- | --- |
| Week 1 | 0.6-0.8 mg/kg/day | BID | 7 days |
| Week 2 | 0.4-0.6 mg/kg/day | BID | 7 days |
| Week 3 | 0.2-0.4 mg/kg/day | BID | 7 days |
| Week 4 | 0 mg/kg/day | BID | 7 days |

## Dosing rationale and calculations

Data on the pharmacokinetics related to cannabinoids in adolescents are extremely lacking. In the current study, the CBD dose is extrapolated from safety data obtained in clinical trials in children and adults with refractory epilepsy [32–38]. CBD was found to be safe and well-tolerated in children with refractory epilepsy at a dose of up to 20 mg/kg/day [32, 33, 35, 38, 39]. We aim to keep the CBD dose as low as possible to limit adverse events and reduce costs for families should we confirm a tolerable dose is effective in future randomized controlled trials.

The use of THC is associated with some risk of developing adverse effects of the central nervous system, [40, 41] however, THC possesses its central pain-relieving potential at very low doses, much lower than would be expected from recreational cannabis exposures [42]. Previous studies on a 1:20 THC:CBD CHE oil in children reported that plasma THC levels following doses of up to 12 mg/kg/day suggested a low risk for THC intoxication [21]. The quantity of THC in our study product is 1 mg per mL with maximum THC doses <u>only reaching 0.05 mg/kg/day</u>, which we do not expect to be associated with any adverse psychoactive reactions. In this study, the maximum dose of CBD will be <u>less than 10 percent</u> of the recommended dose of CBD in the previous studies conducted in the pediatric population [43]. In order to maintain the accuracy and consistency in the dosing regimen of study participants across all the study centers, the mid-point of the dose range will be selected to calculate the desired dose based upon the weight of the participants. Participants will be weighed at each study visit to assist in tracking changes in appetite; however, the dose calculation will be based on the weight taken at baseline. The final dose will be calculated by rounding off (0.5 mL of MPL-001). This will help in achieving improved precision and will ease administration of the investigational product to the study participants. For example, an adolescent weighing 50 kg will receive a starting total daily dose containing 15 mg of CBD (0.3 mg/kg/day) and take 4 mLs in the morning and 11 mLs in the afternoon.

## Dose limiting toxicities (DLTs)

Adverse events will be categorized using the Common Terminology Criteria for Adverse Events (CTCAE version 5.0 dated 27 Nov 2017). If any of the following DLTs occur the participant will not move up to the next dose level. Dose escalations will not be reattempted but the participant shall remain in CAN-CHA should no other DLTs occur. DLTs include:

1. Parental/youth report complaints of moderate mood elevation defined as exaggerated feelings of well-being which is disproportionate to the events and stimuli (Euphoria Grade 2)
2. Somnolence Grade 2 which includes moderate sedation (sleepiness and drowsiness) that limits instrumental activities of daily living
3. Cannabis-attributed diarrhea, Grade 2 or more defined as an increase of 4 – 6 stools per day over baseline; moderate increase in ostomy output compared to baseline; limiting instrumental activities of daily living
4. Unexplained tachycardia (w/out pain, fever, anemia etc.) requiring medical intervention
5. Unexplained hypotension requiring medical intervention
6. Non-infectious conjunctivitis Grade 2 defined as moderate decrease in visual acuity (best corrected visual acuity 20/40 and better or 3 lines or less decreased vision from known baseline) characterized by inflammation, swelling and redness to the conjunctiva of the eye.
7. Serious adverse events requiring hospitalization
8. Discretion of the participant, physician, or parents

## Primary Outcome

The frequency and type of cannabis-related adverse events among study participants will be assessed daily throughout the study. Adverse events will be reported daily and reviewed at study visits.

## Secondary Outcomes

1. The frequency of headache measured using headache-free days [assessed: daily throughout the study]. Reported number of headache-free days per month during the study period
2. The average intensity of pain due to chronic headaches as measured using an 11-point Numeric Rating Scale (NRS) [assessed: daily throughout the study] [44]. Reported for each study as a percentage change in average daily pain intensity due to chronic headaches on the numeric rating scale (NRS) from baseline to each follow up visit
3. The impact of pain on participants’ quality of life using the PROMIS Pediatric Pain Interference– Short Form 8a [assessed: at visits 1,2,3,4,5,6 and 7] [26]. Reported as a percentage change in the scores from baseline value
4. The quality of sleep will be recorded using the PROMIS Pediatric Sleep-Related Impairment– Short Form 8a scale [45]. [assessed: at visits 1,2,3,4,5,6 and 7] Reported as a percentage change in scores from baseline value
5. Changes in anxiety will be measured using the PROMIS Pediatric Short Form v2.0 – Anxiety – 8a scale [46] [assessed: at visits 1,2,3,4,5,6 and 7]. Reported as percentage change in scores from the baseline value
6. Change in mood will be evaluated using two tools PROMIS Pediatric Short Form v2.0 – Depressive Symptoms 8a scale and the PROMIS Pediatric Positive Affect – Short Form 8a. [29, 47] [assessed: at visits 1,2,3,4,5,6 and 7]. Reported as a percentage change in scores from baseline value
7. Change in self-directed goal attainment (participant and parent reported) [assessed: monthly throughout the study] [30]. Reported as a percentage toward a physical, mental and social by participant at each monthly visit
8. Change in scores of PedsQL ™ Family Impact Module, Version 2.0 [assessed: monthly throughout the study] [31]. Reported as percentage change in scores from the baseline value
9. Steady-state trough plasma levels of bioactive cannabinoids THC, CBD, 11-OH-THC, 7-OH-CBD, and endocannabinoids [assessed: monthly throughout the study]. Reported as a plasma concentration relative to each dose increase/decrease and according to genotype
10. Genetic polymorphisms within genes encoding for cytochrome P450 enzymes and the p-glycoprotein transporter and their association with plasma levels of THC, CBD, and their active metabolites in the study participants

## Assessments

Participants will be enrolled in the study for six months. De-identified participant data will be collected electronically in a Research Electronic Data Capture (REDCap) database or paper copies will be provided based on participants’ preferences. Assessments completed on paper will be kept as source documents at each study site and entered into REDCap by study staff. The following includes a detailed description of all planned assessments.

## Physical examination

During the baseline and close out visits (visit 7), participants will undergo physical assessments, but weight will be reported at every visit to potentially signal changes in appetite. Blood work will assess liver function and renal safety, plasma concentration of cannabinoids, and pregnancy status. At the time of screening, participants will receive an electrocardiogram as per standard practice at each site should they not have had one in the previous 3 months.

## Electronic headache diary

The daily diary was developed in collaboration with the youth advisors as described above. Administration will incorporate best practices in the use of daily diaries in adolescents, and feedback from patients with chronic pain and families interviewed about the use of in-home longitudinal ecological momentary assessment tools [48]. The purpose of the diary is to monitor and report headache characteristics, including intensity (Numeric Rating Scale; NRS), headache-free days, use of the rescue medication, and potential headache triggers (e.g., low sleep). Electronic diaries will be completed by the participants using REDCap. Adverse events will be assessed daily with flags to research coordinators to review changes. The NRS is a validated tool that will be used to record daily self-reports of headache intensity from visit 1 until the end of the study [49]. Participants will rate the intensity of pain from 0 to 10, 0 means no hurt, 10 means the worst hurt you could ever imagine. The efficacy of the investigational product in treating the chronic headaches will be assessed in reducing the intensity of headache from baseline to each follow-up visit. Daily data from the electronic headache diaries will be transferred to the server. Based on pain intensity reported on the NRS, the study participant will be categorized into the severity of the headaches. Mild headache will vary in intensity between 1-5, moderate between 6-8, and severe between 9-10 [50]. A complete list of the daily headache diary questions are provided in Table 3. All the study participants will be given an option of using an electronic wrist worn device (actigraphy), provided at no cost to them by the study team, to help keep track of the number of hours slept per night.

**Table 3.**
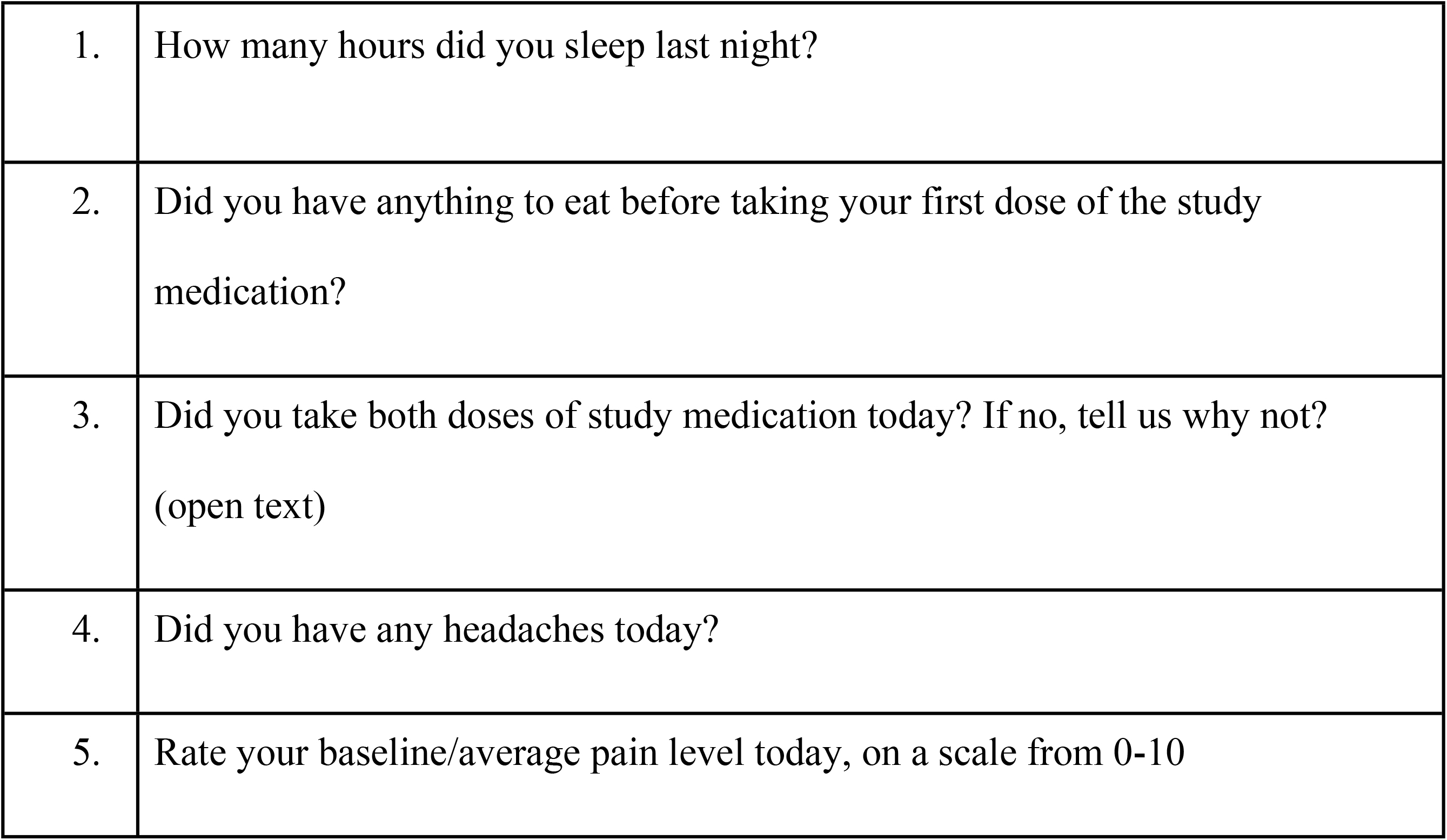
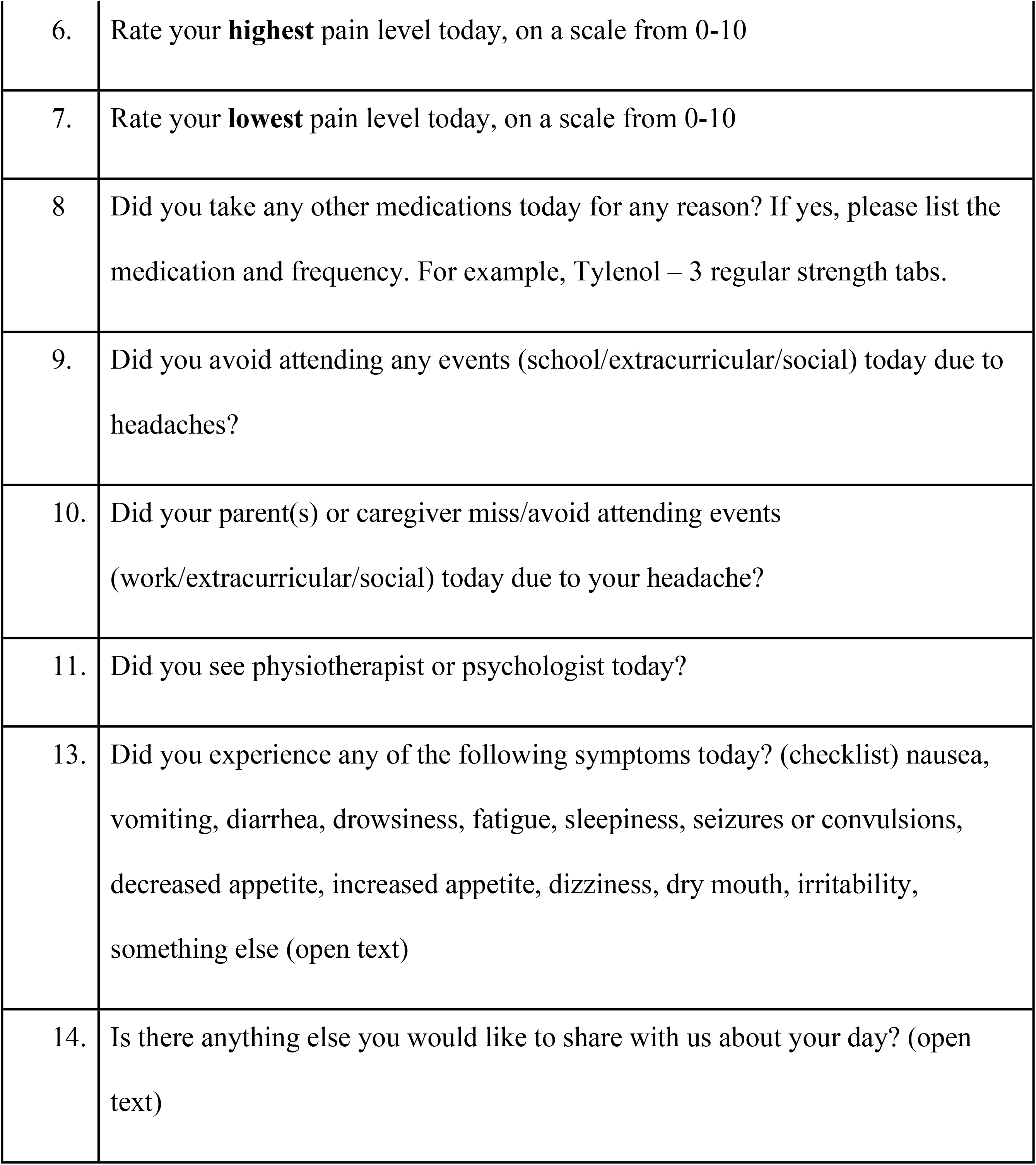
Electronic daily headache diary questions.

|  |  |
| --- | --- |
| 1. | How many hours did you sleep last night? |
| 2. | Did you have anything to eat before taking your first dose of the study medication? |
| 3. | Did you take both doses of study medication today? If no, tell us why not?<br>(open text) |
| 4. | Did you have any headaches today? |
| 5. | Rate your baseline/average pain level today, on a scale from 0-10 |
| 6. | Rate your **highest** pain level today, on a scale from 0-10 |
| 7. | Rate your **lowest** pain level today, on a scale from 0-10 |
| 8 | Did you take any other medications today for any reason? If yes, please list the medication and frequency. For example, Tylenol – 3 regular strength tabs. |
| 9. | Did you avoid attending any events (school/extracurricular/social) today due to headaches? |
| 10. | Did your parent(s) or caregiver miss/avoid attending events (work/extracurricular/social) today due to your headache? |
| 11. | Did you see physiotherapist or psychologist today? |
| 13. | Did you experience any of the following symptoms today? (checklist) nausea, vomiting, diarrhea, drowsiness, fatigue, sleepiness, seizures or convulsions, decreased appetite, increased appetite, dizziness, dry mouth, irritability, something else (open text) |
| 14. | Is there anything else you would like to share with us about your day? (open text) |

## PROMIS measures

Patient-Reported Outcomes Measurement Information System (PROMIS) evaluates social, mental, and physical health of adults and children [51]. We will be using the PROMIS tools to measure pain interference, sleep, depressive symptoms, emotions, and family’s quality of life. These tools will be completed by the study participants. The following PROMIS measure will be administered by the study participants.

## Pediatric Pain Interference – Short Form 8a scale

The Pediatric Pain Interference PROMIS tool assesses the self-reported consequences of pain on the essential aspects of a participant’s life. It measures the extent to which pain interferes with cognitive, emotional, social, physical, and recreational activities. The pediatric pain interference scale is non-disease specific, and the items utilize a 7–day recall period (items include the phrase “the past 7 days”). Varni et al. describe the strong psychometric properties of the PROMIS Pediatric Pain Interference Scale [26].

## Pediatric Sleep-Related Impairment – Short Form 8a scale

The Pediatric Sleep-related Impairment is a PROMIS tool, which assesses perceptions of sleepiness during awake hours and sleep-related impairments during the day. Mainly this tool assesses sleepiness during the daytime, sleep offset, and its impact on cognition, other daily activities, as well as emotional impact. The Pediatric Sleep-Related Impairment – Short Form 8a scale has demonstrated good validity and internal consistency [27].

## Pediatric Depressive Symptoms – Short Form 8a scale v2.0

Pediatric Short Form v2.0 – Depressive Symptoms 8a is a PROMIS tool which is used for assessment of negative mood states, including sadness or guilt, self-criticism, self-perceived worthlessness, loneliness, interpersonal alienation (social cognition) and decreased positive affect and engagement. However, this scale does not assess somatic symptoms associated with depression, such as changes in appetite and sleeping patterns. Pediatric Short Form v2.0 – Depressive Symptoms 8a scale has demonstrated good test-retest reliability, and validity of administration across groups [28].

## Pediatric Positive Affect – Short Form 8a scale

The Pediatric Positive Affect scale is PROMIS tool used for the assessment of positive or rewarding feelings and moods, including elation, pride, affection, pleasure, joy, engagement, excitement, happiness, and contentment. The short form of Pediatric Positive Affect scale is an efficient, accurate, and valid assessment of positive affect in adolescents and good reliability has been reported [47].

## Pediatric Anxiety Symptoms-Short Form 8a scale v.2.0

The pediatric short form v2.0 Anxiety scale is a PROMIS tool used to assess the self-reported fear (fearfulness, panic), anxious misery (worry, dread), hyperarousal (tension, nervousness, restlessness), and somatic symptoms related to arousal (racing heart, dizziness). The literature reports excellent reliability and validity of the Pediatric Short Form v2.0 – Anxiety – 8a scale [29].

## Goal attainment Scaling

Goal attainment scaling is a patient-derived and reported outcome measure in which participants select their goals in areas where they would like to improve function [30]. Participants will be asked to select three goals at their screening appointment which could include physical, social, academic, or personal goals that they hope to improve on throughout the study period. In this, scaling goals are weighted with the importance (on a 0-10 scale) and difficulty (also on a 0-10 scale).

## PedsQL™ Family Impact Module

Pediatric quality of life family impact module aims to measure the effect of the child’s chronic health condition on the family and parents. The PedsQL examines the domains of physical, emotional, social, cognitive, worry and communication to provide an overview of quality of life and impact on families. The PedsQL scale has demonstrated well defined validity and reliability [31].

## Cannabinoid and metabolite assessment

To establish the relationship between dose-escalation and steady-state trough plasma concentrations (Css, Min) of the bioactive cannabinoids, we will collect blood samples before each dose escalation. At the first visit the study nurse will ask participants what methods they would like to employ to reduce needle pain. They will be offered evidence-based strategies including distraction and breathing techniques, use of a topical anesthetic and preferred positioning for blood draws [52]. Cannabinoid levels will be assessed using a previously described Liquid Chromatography-Mass Spectrometry (LC-MS/MS) method validated as per US FDA guidelines [53]. The blood sample from the study participants will be drawn into the heparinized lithium Barricor vacutainers ® (BD Canada, Mississauga, ON). The collected blood will be centrifuged for 10 minutes at 1500 rpm to separate the plasma from the blood. The separated plasma will be stored at − 80 °C until analysis. An LC-MS/MS method will be used to quantify the plasma concentration of THC, CBD, 11-OH-THC, 7-OH-CBD, and Cannabichromene. Stock solutions of cannabinoids and their respective stable isotope-labeled internal standards (Cerilliant Corp., Round Rock, TX) will be prepared in methanol at 1 mg/mL and stored at –20 °C until use. Stock solutions will be serial diluted using blank human plasma to develop calibration curves. Low, medium, and high-quality control (QC) samples will determine acceptance of the analytical run. QC and calibration samples will be prepared fresh each day samples are analyzed. Calibration curves will be constructed from a plot of the peak area ratios of the cannabinoid and its respective internal standard against the nominal concentration. The linearity of the calibration curves will be determined with a linear least-squares regression analysis using 1/X as a weighting factor. Using FDA bioanalytical guidance, all calibration curves will be linear (r2 >0.99) and intra– and inter-day precision and accuracy will be within 15%, except at the limit of quantification which will need to be within 20%. For all QC samples for a given run calculated concentrations will be within 15% of their nominal values. The plasma sample extraction will involve addition of internal standard working solution (10 μL) followed by protein precipitation with cold acetonitrile and removal of lipids using Agilent Captiva EMR-Lipid well plates. Supernatant will be transferred to borosilicate tubes and dried under filtered air for 30 minutes at 37°C. Samples will be reconstituted with 200 mL mobile phase and 100 μL transferred into HPLC inserts, 5 μL will be further injected onto an Agilent Phenyl-hexyl small-bore (2.1 × 12.5 mm, 5 μm) column and guard column maintaining the column temperature at 30°C to separate cannabinoids. Mobile phase A (water containing 0.1 mM ammonium formate) and B (methanol containing 0.1 mM ammonium formate) will separate the cannabinoids using a 10 minute gradient method with a flow rate of 250 L/min. Positive ion mode electrospray ionization (ESI), with an ion spray voltage of 5500 V, will be used to conduct multiple reaction monitoring (MRM) for quantification of CBD (quantifier ion m/z 315.1>193.2; qualifier ion m/z 315.1>259.2), THC (quantifier ion m/z 315.1>193.1; qualifier ion m/z 315.1>259.2), 11-OH-THC (quantifier ion m/z 331.1>193.1; qualifier ion m/z 331.1>201.0), CBC (quantifier ion m/z 315.1>193.2; qualifier ion m/z 315.1>259.2), CBD-d3 (318.2>196.1), THC-d3 (318.2>196.1), 11-OH-THC-d3 (334.2>196.1), and CBC-d9 (324.3>202.2) using a ABSciex 6500 QTRAP mass spectrometer (ABSciex, Concord, ON, Canada) and MultiQuant 3.0.1 Software. The temperature, gas source 1, gas source 2, curtain gas, entrance potential, and collision activated dissociation gas were set to 600°C, 70, 60, 50, 10 V, and 10, respectively.

The experienced team at the Cannabis Research Initiative of Saskatchewan, University of Saskatchewan will conduct all the analyses of the samples collected from all the three study sites. The samples will be analyzed in the three batches starting within the three months of the collections. The stability analysis of cannabinoids stored at –80°C for three months indicates excellent stability as described previously [20].

## Pharmacogenomic analyses

Genomic DNA will be extracted from saliva samples using a QIAmp DNA extraction kit at the University of Manitoba and genotyping will be performed using the Illumina Global Screening Array (GSA) v3 array on the iScan array scanner. The GSAv3 GWAS arrays have been designed to capture both common and rare genetic variation, including all variants that have been shown by the Clinical Pharmacogenetics Implementation Consortium to affect the function of the selected candidate genes [54]. Genotyped variants and samples will undergo standard quality control procedures in PLINK 1.9 [55]. Imputation of non-genotyped variants will be performed using the Michigan Imputation Server, including the Haplotype Reference Consortium data as a reference and genetically-determined ancestry will be assessed with EIGENSOFT v5 [56]. Association between these variants and response outcomes will be performed using logistic regression.

## Health care utilization

Costs are an important consideration for families caring for an adolescent with a chronic illness, and are especially concerning for families accessing cannabis oils [57]. Participants in this study will provide consent to access their administrative health records for a pre and post study of healthcare utilization to be designed, reviewed and approved in the future. During CAN-CHA, measures of absences from school and work for participants and caregivers, hospitalizations and the use of rescue medicines will be captured to inform building blocks for future health technology assessment. To reduce participant burden, access to health records will also ensure that the use of all rescue medications is documented should there be an admission to the emergency department or inpatient unit.

## Sample size

CAN-CHA is a tolerability study, designed to evaluate the safety of escalating doses of a cannabidiol-enriched CHE. Given the within-participant study design, a sample size of 20 study participants (common for early phase trials) should provide a reasonable characterization of the pattern of adverse event frequency and severity as dosing increases. To provide more generalizable data we will recruit these participants across three pediatric chronic pain/headache programs in Halifax, Vancouver, and Toronto, Canada.

## Data collection, management and sharing

Data collection will be the responsibility of the study team at each site under the supervision of the site investigators (TFO, ECL, GAF) and trial sponsor (LEK, University of Manitoba).

Investigators and research coordinators will be responsible for ensuring the accuracy, completeness, legibility, and timeliness of the data reported. All source documents will be completed in a neat, legible manner to ensure accurate interpretation of data. Hardcopies of the study visit measurement tool will be provided as source documents for each participant enrolled in the study. Data recorded in the electronic case report form (eCRF) derived from source documents should be consistent with source documents. Clinical and laboratory data will be entered into REDCap (Research Electronic Data Capture), [58] a 21 CFR Part 11-compliant data capture system provided by the Women and Children’s Health Research Institute at the University of Alberta, Edmonton. REDCap includes password protection and internal data quality checks, such as automatic range checking, to identify data that appear inconsistent, incomplete, or inaccurate to the study team for verification. Clinical data will be entered directly from the source documents. Full de-identified datasets will be available from the corresponding author upon reasonable request and review by the trial steering committee.

## Statistical analysis

The primary statistical analyses will be a descriptive summary of the pattern of incidence and severity of adverse events across dosage levels. Adverse event severity grading is assigned using CTCAE definitions as described above. Conventional summary statistics will be used to describe baseline characteristics and other outcomes (means, standard deviations, as well as medians, range, and interquartile range (IQR) for numerical variables; counts and percentages for categorical variables). Adverse events will be reported overall (duration of the study period) and by dosage level (study month). Medians, ranges and IQR will be provided for the concentrations of CBD, THC, and the major metabolites at each sampling point. The ratio of concentration of parent compound to metabolites and endocannabinoids will also be summarized to explore variability in cannabinoid metabolism. Severity, frequency, and relationship of treatment emergent AEs to study intervention will be presented by system organ class and MEDRa coded. All serious adverse events will be reported to pharmacovigilance programs including Health Canada and the Canadian Paediatric Surveillance Program along with regulatory bodies including research ethics boards, the data safety monitoring board and trial steering committee. The secondary outcomes including pain, sleep impairment, depression, positive affect, anxiety, and goal attainment scores will be summarized at each timepoint; within-participant change from baseline of these measures will also be summarized (both absolute and percentage change) but this trial is underpowered, and not designed to evaluate efficacy.

## Ethics and dissemination

CAN-CHA received a No Objection Letter from Health Canada (Dec 2022), registered with ClinicalTrials.gov (NCT05337033). The study was approved by the University of Manitoba Health Research Ethics Board (HS25503-B2022:037. An institutional cannabis research license is received in Sep 2022, and we expect to enroll our first participant in Spring 2023. Written informed consent will be taken from all the participants and from the legal guardians for the participants who will be below 16 years of age. We plan to disseminate our findings of the CAN-CHA trial by presenting them to conferences and publishing them in an open-access journal.

## Discussion

In Canada, adolescents with refractory headache disorders are using cannabis products off-label to manage their symptoms, self-guided in the absence of evidence [59]. Chronic headaches are often resistant to standard drug therapies in adolescents, resulting in school absenteeism, withdrawal from social activities and can cause significant stress for families [60]. Based on anecdotal reports, parents of children with chronic headaches found cannabinoids to be effective for the management of headache[1], but there remains minimal data available to inform dosing or safety. CAN-CHA is a tolerability study designed with youth and parents that will investigate the safety of escalating doses up to 1mg/kg/day of CBD-enriched CHE in adolescents. This represents the first study evaluating a cannabis product, providing valuable knowledge on the safety and dosing of a CBD-enriched CHE in children with chronic pain. The CAN-CHA trial is an open label with a small sample size, underpowered to evaluate efficacy. This study will inform dose selection for a larger scale randomized clinical trial comparing a CBD-enriched CHE oil to placebo (on top of standard of care) for adolescents with chronic headache. Given the high prevalence of chronic headaches, severe morbidity, and current lack of effective treatment options, the successful outcomes from this research project will have the potential to create meaningful impact for the lives of Canadian adolescents.

## Supporting information

**S1.** SPIRIT checklist: Recommended items to address in a clinical trial protocol and related documents.

**S2.** Approval by ethics committee.

**S3**. Study protocol reviewed by ethics committee.

## Abbreviations

AE: Adverse Events; BMI: Body Mass Index; CBD: Cannabidiol; CHE: Cannabis Herbal Extract; CNS: Central Nervous System; eCRF: electronic case report form; GMP: Good Manufacturing Practices; NRS: Numeric Rating Scale; NSAIDs: Non-steroidal Anti-inflammatory Drugs; REDCap: Research Electronic Data Capture; THC: delta 9 – Tetrahydrocannabinol

## Ethics and dissemination

The study was approved by the University of Manitoba Health Research Ethics Board (HS25503-B2022:037). We plan to disseminate our findings of the CAN-CHA trial open-access publication.

## Author’s contributions

LEK is the principal Investigator; led the proposal and protocol development. ECL, GAF, and TFO are the qualified investigators contributed to study design and to development of the proposal. TL, ZA, MCG, are the youth research partners, RB is biostatistician, BS is trial coordinator, and MC is PhD trainee; drafted and revised manuscript. All authors provided insights in study design, read, and approved the final manuscript.

## Availability of data and material

Access to the protocol and a deidentified dataset may be provided upon reasonable request to **LEK** and approval by the Clinical Trial Steering Committee.

## Data Availability

Deidentified research data will be made publicly available when the study is completed and published.

## Acknowledgements

We are thankful to Dr Thierry Lacaze-Masmoniteil for providing insights during the design of the study and all of the C4T members who have been consulted on protocol development and mentorship.

